# Benefits of Aeroallergen Testing on Oral Corticosteroid Bursts in Adults with Asthma

**DOI:** 10.1101/2024.01.29.24301962

**Authors:** Patrick K. Gleeson, Knashawn H. Morales, Timothy M. Buckey, Olajumoke O. Fadugba, Andrea J. Apter, Jason D. Christie, Blanca E. Himes

## Abstract

**BACKGROUND:** Aeroallergen testing can improve precision care for persistent asthma and is recommended by the U.S. clinical guidelines. How testing benefits diverse populations of adults with asthma, and the importance of the testing modality used, are not fully understood.

**OBJECTIVE:** We sought to evaluate whether receipt of aeroallergen testing was associated with a reduction in oral corticosteroid (OCS) bursts.

**METHODS:** We used electronic health record data to conduct a retrospective, observational cohort study of adults with asthma who were prescribed an inhaled corticosteroid and had an Allergy/Immunology visit in a large health system between 1/1/2017-6/30/2022. Negative binomial regression models were used to evaluate whether OCS bursts in the 12-month period after an initial visit were reduced for patients who received aeroallergen testing. We also measured differences in benefit after excluding patients with chronic obstructive pulmonary disease (COPD) and smoking histories, and whether testing receipt was via skin prick or serum.

**RESULTS:** 668/1,383 (48.3%) patients received testing. Receipt of testing was not associated with fewer bursts in all patients (incidence rate ratio (IRR)=0.83 versus no testing, p=0.059), but it was among never smokers without COPD (417/844 tested, IRR=0.68, p=0.004). The receipt of *skin* testing was associated with fewer bursts in all patients (418/1,383 tested, IRR=0.77, p=0.02) and among never smokers without COPD (283/844 tested, IRR=0.59 versus no testing, p=0.001).

**CONCLUSION:** Guideline-concordant aeroallergen testing in the context of Allergy/Immunology care was associated with clinical benefit in a real-life, diverse cohort of adults with asthma. This benefit varied according to patient comorbidities and the testing modality.

## INTRODUCTION

Asthma is a common chronic disease in adults and a major public health burden in the U.S.^1–3^ A key component of asthma care is to reduce symptoms and exacerbations by addressing modifiable risk factors, including aeroallergen exposure in sensitized patients.^2,4^ Pollens, dust mite, cat, dog, mold, cockroach, and mouse are common aeroallergens and important asthma triggers.^5–19^ Addressing allergic asthma triggers is beneficial in adults with asthma, as most are sensitized to one or more aeroallergens and aeroallergen exposure is nearly universal in U.S. homes.^20–23^ Patient history alone is unreliable for determining allergic sensitization.^4,24^ In randomized controlled trials of adults with asthma, aeroallergen testing with allergen avoidance education improved patients’ awareness of perceived asthma triggers and lung function^25^ and complemented asthma self-management.^26^ National Heart, Lung, and Blood Institute (NHLBI)-led online focus groups of people with asthma that explored patients’ values and priorities related to their asthma found that most patients took steps to mitigate indoor allergen exposure.^27^ Thus, testing aligns with patients’ strategies to reduce asthma symptoms.

The U.S. clinical guidelines recommend aeroallergen testing for patients with persistent asthma, defined as asthma which requires Step 2 treatment or higher to maintain control, and the preferred controller treatment at Step 2 is inhaled corticosteroid (ICS)-containing treatment.^4,27^ However, allergen mitigation may be difficult or costly, and the benefit of testing in large numbers of real-life adults with asthma is not well-studied. In prior work, we found that among 1,789 adults with asthma who established outpatient care in a large health system, were prescribed an ICS, and received aeroallergen testing, oral corticosteroid (OCS) bursts were reduced in the 12-month period after testing compared with before.^20^ Consistent with the clinical focus of providers, testing was more often performed in the context of asthma subspecialty clinics, specifically Allergy/Immunology and Pulmonary rather than Primary Care (odds ratios (OR) of 91.3 and 7.1 versus Primary Care, respectively). However, that study was unable to determine whether improved outcomes were due to allergy testing and test-based management versus other interventions that may have occurred during specialist visits. Moreover, although many patients with asthma have smoking histories and/or comorbid COPD, and such asthma is often refractory to standard care strategies, the two clinical trials that evaluated the benefits of aeroallergen testing limited or excluded these patients.^25,26^ Finally, prior investigations have not evaluated the relative benefits of skin prick or serum-specific immunoglobulin E (IgE) testing among adults with asthma. Because skin prick testing is only performed in practices with skin testing supplies and trained staff (e.g., Allergy/Immunology clinics), whereas serum-specific IgE testing can be ordered by any provider, the benefits of each modality have implications for health systems.

In this study, we sought to determine whether, among adults with asthma who had an Allergy/Immunology visit, the receipt of guideline-concordant aeroallergen testing was associated with fewer OCS bursts after the visit compared with patients who did not receive testing. We then considered whether the effect of testing differed for patients who were never smokers and without comorbid COPD; and whether the reduction in OCS bursts varied according to the receipt of skin prick or serum-specific IgE testing.

## METHODS

### Study design

We conducted a retrospective, observational cohort study of adults with asthma using EHR data from Penn Medicine, a large, diverse health system that serves the greater Philadelphia area, from encounters dated 1/1/2015-6/30/2023. The University of Pennsylvania Institutional Review Board approved our study.

### Study population

We obtained patient-level data and clinical notes for adults (i.e., age ≥ 18 years) who had at least one encounter with an International Classification of Diseases (ICD)-10 code for asthma (J45*) in any of their records; received an ICS prescription from 1/1/2017-6/30/2023; and had two or more outpatient Allergy/Immunology visits at Penn’s primary Allergy/Immunology clinic, for which the first (i.e., index) visit occurred on or prior to 6/30/2022. We chose a two-visit threshold to ensure that all patients had established specialist care and had an opportunity to receive testing. Patient demographic and comorbidity data included age at the index Allergy/Immunology visit (categorized into five levels); gender; race (using EHR categories); ethnicity (Hispanic or Latino status); insurance status (commercial, Medicaid, or Medicare); body mass index (BMI in kg/m^2^, categorized into the five levels used by the Centers for Disease Control and Prevention^28^); smoking category (never, former, or current); COPD (any ICD-10 code of J41*, J42*, J43*, or J44*); and chronic rhinitis (any ICD-10 code of J30* or J31*).

### Timeline of observations included

Figure 1 displays timelines of data considered. We chose 1/1/2017 as the date of the earliest index visit possible because in our health system, emergency department (ED) and hospitalization encounter data from some hospitals in 2015 are missing from the data warehouse, whereas encounter data from 1/1/2016 onward was complete.

**Figure 1.**
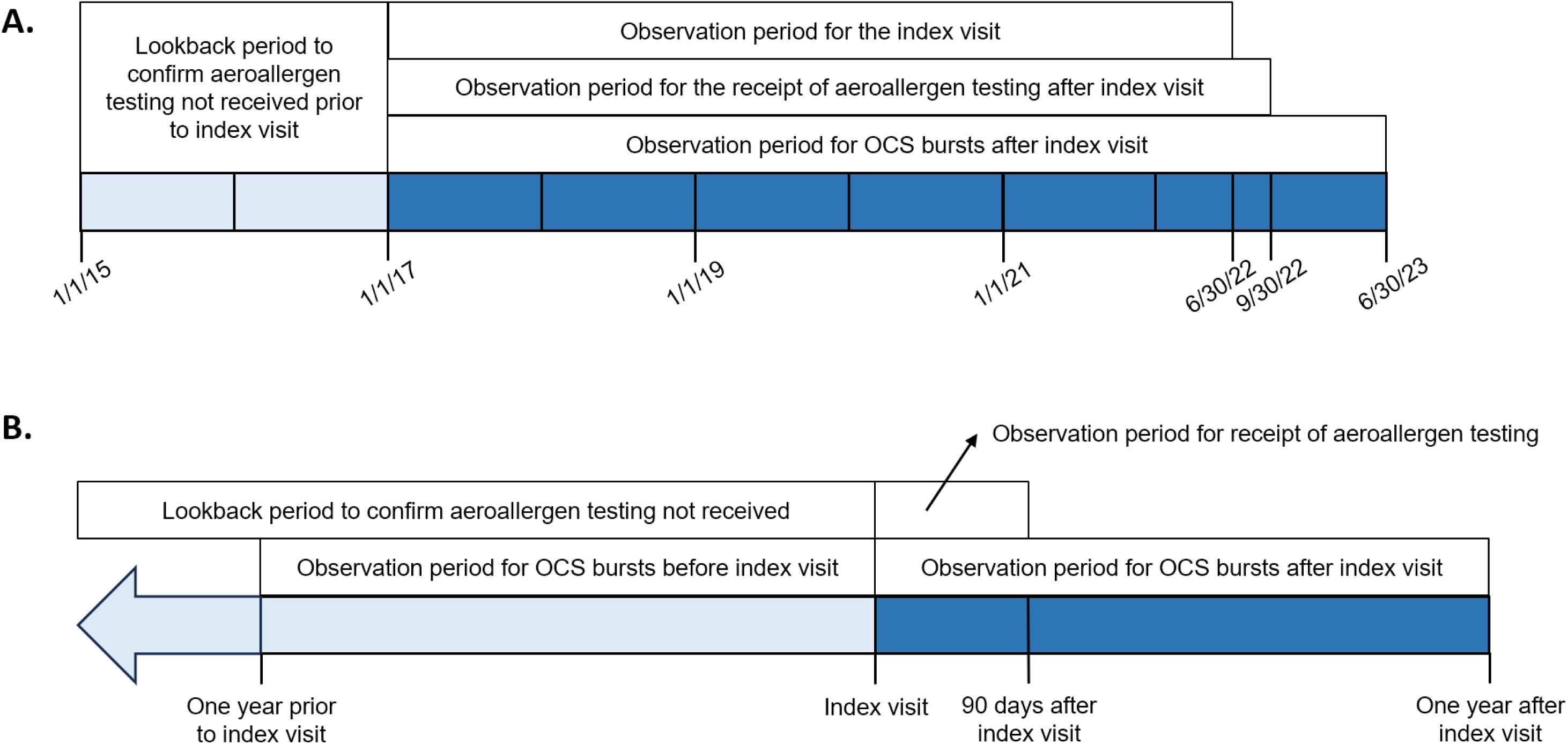
A) Overall Study Period. Shown are the observation periods for the index visit, receipt of aeroallergen testing after the index visit, counting of oral corticosteroid bursts after the index visit, and an additional lookback period to confirm that aeroallergen testing was not received. B) Individual-Level Study Period. This timeline is centered on the index visit date for each patient. Shown are the 90-day observation period for the receipt of aeroallergen testing after the index visit, the 12-month observation periods before and after the index visit for the counting of oral corticosteroid bursts, and the lookback period to confirm that aeroallergen testing was not received.

### Asthma exacerbation measures

We selected three variables to represent asthma exacerbations during the 12-month periods before and after the index visit: OCS bursts, ED visits, and hospitalizations. OCS bursts were categorized into four groups: counts of 0, 1, 2-3, and 4 or more; and ED visits and hospitalizations were each categorized into two groups: counts of 0, and 1 or more. Definitions for these exacerbation measures are described in the Online Repository Text and lists of ICD-10 codes and chief complaints that were used to deem ED visits and hospitalizations as asthma-related are provided in Table E1 of the Online Repository.

### Aeroallergen test data

We defined aeroallergen testing as any skin prick or serum-specific IgE tests to tree, grass, ragweed, dust mite, cat, dog, mold, cockroach, or rodent that were recorded in the EHR as structured data, as previously described.^20^

We collected allergy test data from 1/1/2015 because starting in 2015, most serum-specific IgE tests to aeroallergens at Penn Medicine were performed at an in-house laboratory, and hence, there was standard documentation in place and few missing data. More details are in the Online Repository Text.

### Allergy testing exposure variables

We chose a 90-day threshold for the receipt of testing after the index visit to allow testing to occur within the 3-month follow-up interval that is consistent with guideline care.^2,4^ We considered two definitions of testing after the index visit. The first was a binary variable of no testing or any testing. The second was a three-level categorical variable of 1) no testing, 2) any skin prick testing (i.e., with or without serum-specific IgE testing), or 3) only serum-specific IgE testing. We excluded patients who 1) had structured data indicating that aeroallergen testing was performed from 1/1/2015 until the index visit date, as these patients did not have new testing after the index visit; and 2) had aeroallergen testing performed beyond 90 days of the index visit, as these tests were assumed to be unrelated to the index visit.

### Controller inhaler data

We identified inhaler prescription data from 1/1/2017-6/30/2023 and categorized these into three groups: 1) ICS prescription(s) only, 2) any ICS/long-acting beta-agonist (LABA) prescription(s), and 3) any ICS/LABA plus a long-acting muscarinic antagonist (LAMA) prescription(s).

### Documentation of the reasons that aeroallergen testing was not performed

An Allergy/Immunology specialist performed EHR chart review of 50 charts, selected at random, of patients categorized as not receiving testing according to the structured EHR data to identify the reasons that testing was not performed. The purpose was to understand why Allergy/Immunology providers may have chosen not to perform testing or whether providers had ordered or planned testing that was not performed. The specialist reviewed structured testing data dated earlier than 1/1/2015, external lab results imported into the EHR on or prior to the index visit, and free text in Allergy/Immunology notes on or prior to the index visit. We identified seven *a priori* reasons for why testing was not performed, as detailed in the Online Repository Text, and assigned a reason to each patient. A second reviewer who was an Allergy/Immunology fellow reviewed the same 50 charts to assess the interrater reliability. For any disagreements, the two reviewers attempted to reach consensus.

### Statistical Analysis

STATA version 16.1 was used. We evaluated bivariate associations of patient demographic factors with the receipt of testing versus not receiving testing using Pearson’s chi-squared tests. For the primary analysis, we created a multivariable negative binomial regression model with OCS bursts in the 12-month period after the index visit as the outcome, the binary testing variable as the exposure, and with patient demographic factors (e.g., age category, gender, race, and ethnicity), insurance status, BMI, smoking category, COPD, rhinitis, controller inhaler category, and the exacerbation measures before the index visit included as independent variables. We assessed collinearity among independent variables by computing their variance inflation factors (VIFs). For the outcomes of asthma ED visits and hospitalizations after the index visit, the low numbers for these outcomes precluded meaningful multivariable analysis.

To account for changes in healthcare utilization that occurred during the COVID-19 pandemic, we repeated the model creation while restricting the index visit and 90-day window for testing to occur earlier than Philadelphia’s COVID-19 lockdown date of 3/17/2020.

To evaluate whether patients with asthma who did not have COPD and were never smokers had similar trends in OCS use after receipt of testing, we repeated the primary analysis of the full timeline after 1) excluding patients with COPD and 2) additionally excluding current or former smokers.

Among patients who received testing, we evaluated the bivariate associations of patient demographic factors with the receipt of any skin prick testing versus only serum-specific IgE testing using Pearson’s chi-squared tests. We repeated each of the four primary and secondary analyses using the three-level testing variable to separately evaluate the effects of skin prick and serum-specific IgE testing on OCS bursts. For these analyses, we compared the effect estimates for skin prick and serum-specific IgE testing using postestimation Wald tests.

## RESULTS

### Patient characteristics

Of 1,383 adults with asthma who established care in an Allergy/Immunology clinic and appeared to be eligible for testing according to guidelines, 668 (48.3%) received testing within 90 days of their index visits, and 715 (51.7%) did not receive testing. Compared with patients who did not receive testing, patients who received testing were more likely to be under age 45 years (p<0.001), of Black race (p<0.001), and of Hispanic or Latino ethnicity (p=0.02); to not have Medicare insurance (p<0.001); and to have rhinitis (p<0.001) [Table 1].

**Table 1.**
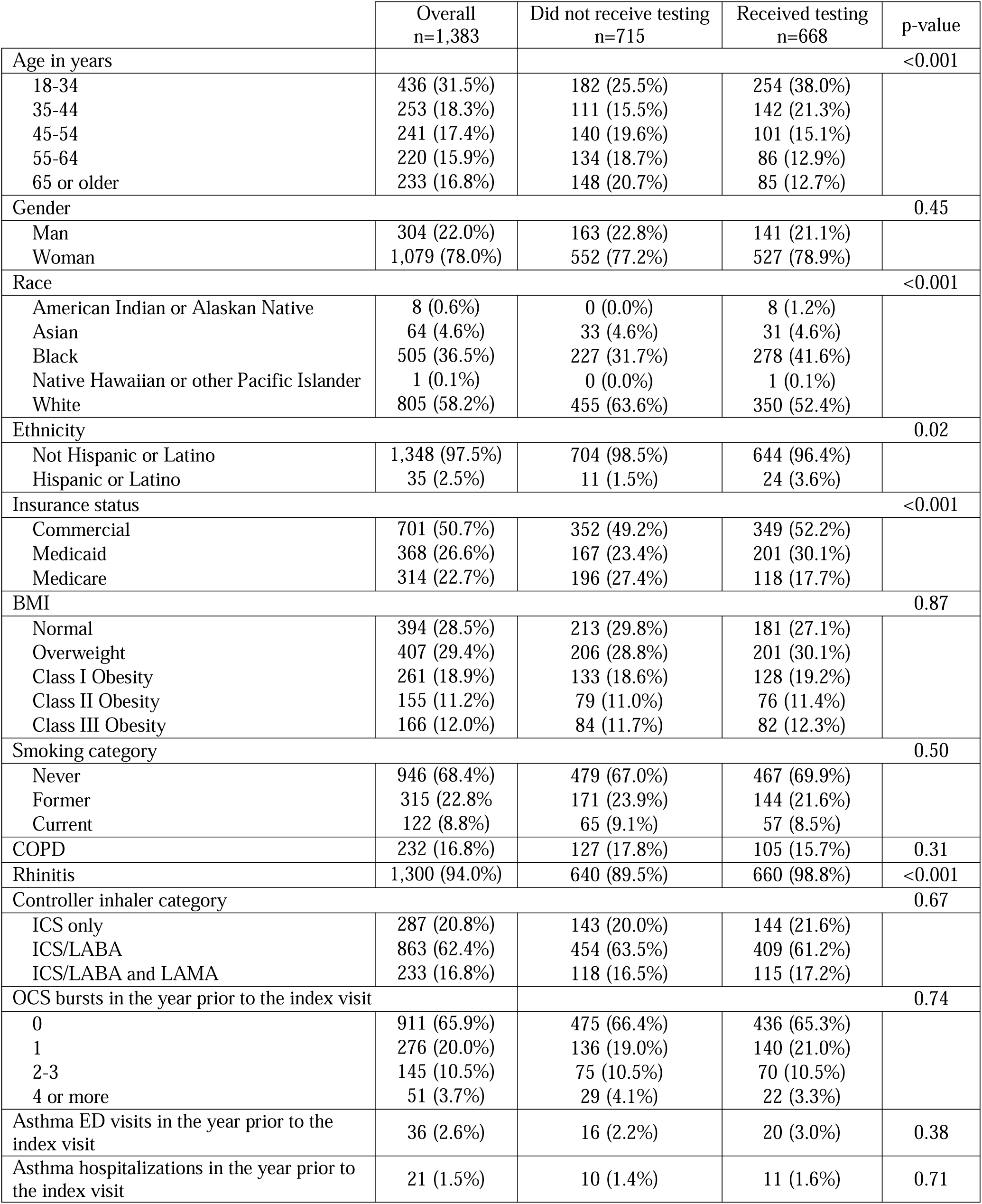
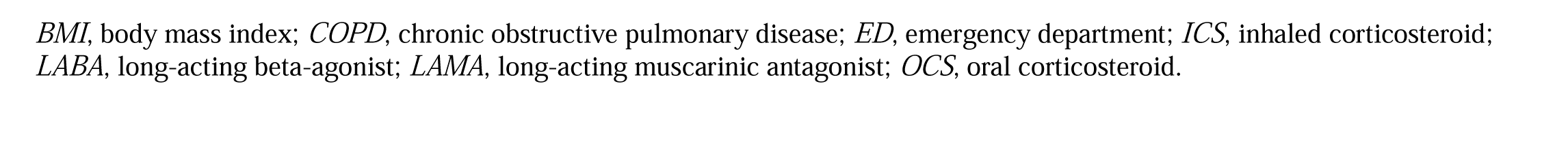
Patient characteristics related to receiving aeroallergen testing among 1,383 adults with asthma. Patients who did not receive any testing (n=715) were compared with patients who received any skin prick or serum-specific immunoglobulin E testing on or within 90 days of an index Allergy/Immunology visit (n=668) using chi-squared tests.

|  | Overall<br>n=1,383 | Did not receive testing<br>n=715 | Received testing<br>n=668 | p-value |
| --- | --- | --- | --- | --- |
| Age in years |  |  |  | <0.001 |
| 18-34 | 436 (31.5%) | 182 (25.5%) | 254 (38.0%) |  |
| 35-44 | 253 (18.3%) | 111 (15.5%) | 142 (21.3%) |  |
| 45-54 | 241 (17.4%) | 140 (19.6%) | 101 (15.1%) |  |
| 55-64 | 220 (15.9%) | 134 (18.7%) | 86 (12.9%) |  |
| 65 or older | 233 (16.8%) | 148 (20.7%) | 85 (12.7%) |  |
| Gender |  |  |  | 0.45 |
| Man | 304 (22.0%) | 163 (22.8%) | 141 (21.1%) |  |
| Woman | 1,079 (78.0%) | 552 (77.2%) | 527 (78.9%) |  |
| Race |  |  |  | <0.001 |
| American Indian or Alaskan Native | 8 (0.6%) | 0 (0.0%) | 8 (1.2%) |  |
| Asian | 64 (4.6%) | 33 (4.6%) | 31 (4.6%) |  |
| Black | 505 (36.5%) | 227 (31.7%) | 278 (41.6%) |  |
| Native Hawaiian or other Pacific Islander | 1 (0.1%) | 0 (0.0%) | 1 (0.1%) |  |
| White | 805 (58.2%) | 455 (63.6%) | 350 (52.4%) |  |
| Ethnicity |  |  |  | 0.02 |
| Not Hispanic or Latino | 1,348 (97.5%) | 704 (98.5%) | 644 (96.4%) |  |
| Hispanic or Latino | 35 (2.5%) | 11 (1.5%) | 24 (3.6%) |  |
| Insurance status |  |  |  | <0.001 |
| Commercial | 701 (50.7%) | 352 (49.2%) | 349 (52.2%) |  |
| Medicaid | 368 (26.6%) | 167 (23.4%) | 201 (30.1%) |  |
| Medicare | 314 (22.7%) | 196 (27.4%) | 118 (17.7%) |  |
| BMI |  |  |  | 0.87 |
| Normal | 394 (28.5%) | 213 (29.8%) | 181 (27.1%) |  |
| Overweight | 407 (29.4%) | 206 (28.8%) | 201 (30.1%) |  |
| Class I Obesity | 261 (18.9%) | 133 (18.6%) | 128 (19.2%) |  |
| Class II Obesity | 155 (11.2%) | 79 (11.0%) | 76 (11.4%) |  |
| Class III Obesity | 166 (12.0%) | 84 (11.7%) | 82 (12.3%) |  |
| Smoking category |  |  |  | 0.50 |
| Never | 946 (68.4%) | 479 (67.0%) | 467 (69.9%) |  |
| Former | 315 (22.8%) | 171 (23.9%) | 144 (21.6%) |  |
| Current | 122 (8.8%) | 65 (9.1%) | 57 (8.5%) |  |
| COPD | 232 (16.8%) | 127 (17.8%) | 105 (15.7%) | 0.31 |
| Rhinitis | 1,300 (94.0%) | 640 (89.5%) | 660 (98.8%) | <0.001 |
| Controller inhaler category |  |  |  | 0.67 |
| ICS only | 287 (20.8%) | 143 (20.0%) | 144 (21.6%) |  |
| ICS/LABA | 863 (62.4%) | 454 (63.5%) | 409 (61.2%) |  |
| ICS/LABA and LAMA | 233 (16.8%) | 118 (16.5%) | 115 (17.2%) |  |
| OCS bursts in the year prior to the index visit |  |  |  | 0.74 |
| 0 | 911 (65.9%) | 475 (66.4%) | 436 (65.3%) |  |
| 1 | 276 (20.0%) | 136 (19.0%) | 140 (21.0%) |  |
| 2-3 | 145 (10.5%) | 75 (10.5%) | 70 (10.5%) |  |
| 4 or more | 51 (3.7%) | 29 (4.1%) | 22 (3.3%) |  |
| Asthma ED visits in the year prior to the index visit | 36 (2.6%) | 16 (2.2%) | 20 (3.0%) | 0.38 |
| Asthma hospitalizations in the year prior to the index visit | 21 (1.5%) | 10 (1.4%) | 11 (1.6%) | 0.71 |

*BMI*, body mass index; *COPD*, chronic obstructive pulmonary disease; *ED*, emergency department; *ICS*, inhaled corticosteroid; *LABA*, long-acting beta-agonist; *LAMA*, long-acting muscarinic antagonist; *OCS*, oral corticosteroid.

### Reasons that aeroallergen testing was not performed

The chart reviewers had 84% agreement in identifying the reasons that testing was not performed. After conferring on 8/50 (16%) charts for whom the reviewers disagreed, consensus was achieved for all charts, and the counts for each reason are shown in Table 2. 19 (38%) patients had prior test results in their charts, and most (n=10) received testing 2-5 years prior to the index visit. Asthma or chronic respiratory disease were listed as secondary problems for 9 (18%) patients, and testing was ordered or planned but not completed for an additional 10 (20%) patients.

**Table 2.** Reasons why aeroallergen testing was not performed on or prior to the index visit according to chart review of 50 charts of patients who did not have codified aeroallergen testing according to structured electronic health record data. Seven reasons are shown in bold along with the number of patients and corresponding percentages, and the reasons are ordered by decreasing percentage.

| Reason for Lack of Aeroallergen Testing According to Chart Review | N (%) |
| --- | --- |
|  | 50 (100%) |
| **Prior test results available in the chart** | 19 (38%) |
| Testing between 2-5 years of index visit | 10 (20%) |
| Testing greater than 5 years before index visit | 4 (8%) |
| Year of testing not specified | 3 (6%) |
| Testing within 2 years of index visit | 2 (4%) |
| **Testing ordered or planned by Penn Allergy/Immunology provider but not completed** | 10 (20%) |
| **Asthma and/or chronic respiratory disease were secondary problems** | 9 (18%) |
| **Patient self-report of previous testing** | 6 (12%) |
| **Other or unspecified reason for testing not performed** | 4 (8%) |
| **Skin prick allergy testing contraindicated** | 1 (2%) |
| **Patient declined testing for any reason** | 1 (2%) |

### Receipt of aeroallergen testing was not associated with change in OCS bursts after the index visit among all patients

According to the primary analysis of 1,383 patients, the receipt of testing trended towards fewer bursts but was not statistically significant (incidence rate ratio (IRR)=0.83 versus no testing, p=0.059) [Table 3]. The IRRs for the remaining independent variables are shown in Table E3 of the Online Repository. The VIFs were less than 1.7, indicating minimal collinearity (Table E4 of the Online Repository). Among 668 patients who received testing, the number of asthma ED visits was 22 before the index visits and 15 after (32% reduction); and the number of asthma hospitalizations was 16 before and 10 after (38% reduction). In contrast, among 715 patients who did not receive testing, the number of ED visits was 18 before index visit and 15 after (17% reduction); and there was no reduction in hospitalizations.

**Table 3.** The effect of aeroallergen testing on oral corticosteroid burst count in the 12-month period after an index Allergy/Immunology visit. The IRRs and p-values were adjusted for all confounding variables in the multivariable models using data from the full cohort of 1,383 adults with asthma who established outpatient care with Allergy/Immunology. This model was repeated for subgroups of the 1,383 patients: 973 whose index visit and post-visit 90-day period occurred prior to the COVID-19 lockdown; 1,151 who did not have COPD; and 844 who did not have COPD and were never smokers.

|  | Overall cohort<br>(n=1,383) |  | Index visit prior to COVID-19 lockdown<br>(n=973) |  | Overall cohort, no COPD<br>(n=1,151) |  | Overall cohort, no COPD, never smokers<br>(n=844) |  |
| --- | --- | --- | --- | --- | --- | --- | --- | --- |
|  | IRR (95% CI) | p-value | IRR (95% CI) | p-value | IRR (95% CI) | p-value | IRR (95% CI) | p-value |
| Aeroallergen testing (none as reference) | 0.83 (0.69, 1.01) | 0.059 | 0.71 (0.55, 0.92) | 0.01 | 0.80 (0.64, 0.99) | 0.042 | 0.68 (0.52, 0.88) | 0.004 |

*COPD*, chronic obstructive pulmonary disease; *IRR*, incidence rate ratio.

### Influence of the COVID-19 pandemic

Figure 2 shows the rates of skin prick and serum-specific IgE tests over time in 3-month intervals. From 2017 to April 2020, skin prick testing was performed at a higher rate than serum-specific IgE testing for except October-December 2019, when the rates were the same. After the COVID-19 lockdown began in March 2020, the skin prick testing rate fell to its nadir during April to June 2020, whereas the serum-specific IgE testing rate increased from the previous interval. Thereafter, the skin prick testing rate increased, and the rates of each test modality were similar for most subsequent intervals. 973/1,383 (70.4%) index visits occurred prior to the COVID-19 lockdown date, and for these patients, the receipt of testing was associated with fewer OCS bursts (IRR=0.71, p=0.01) [Table 3].

**Figure 2.**
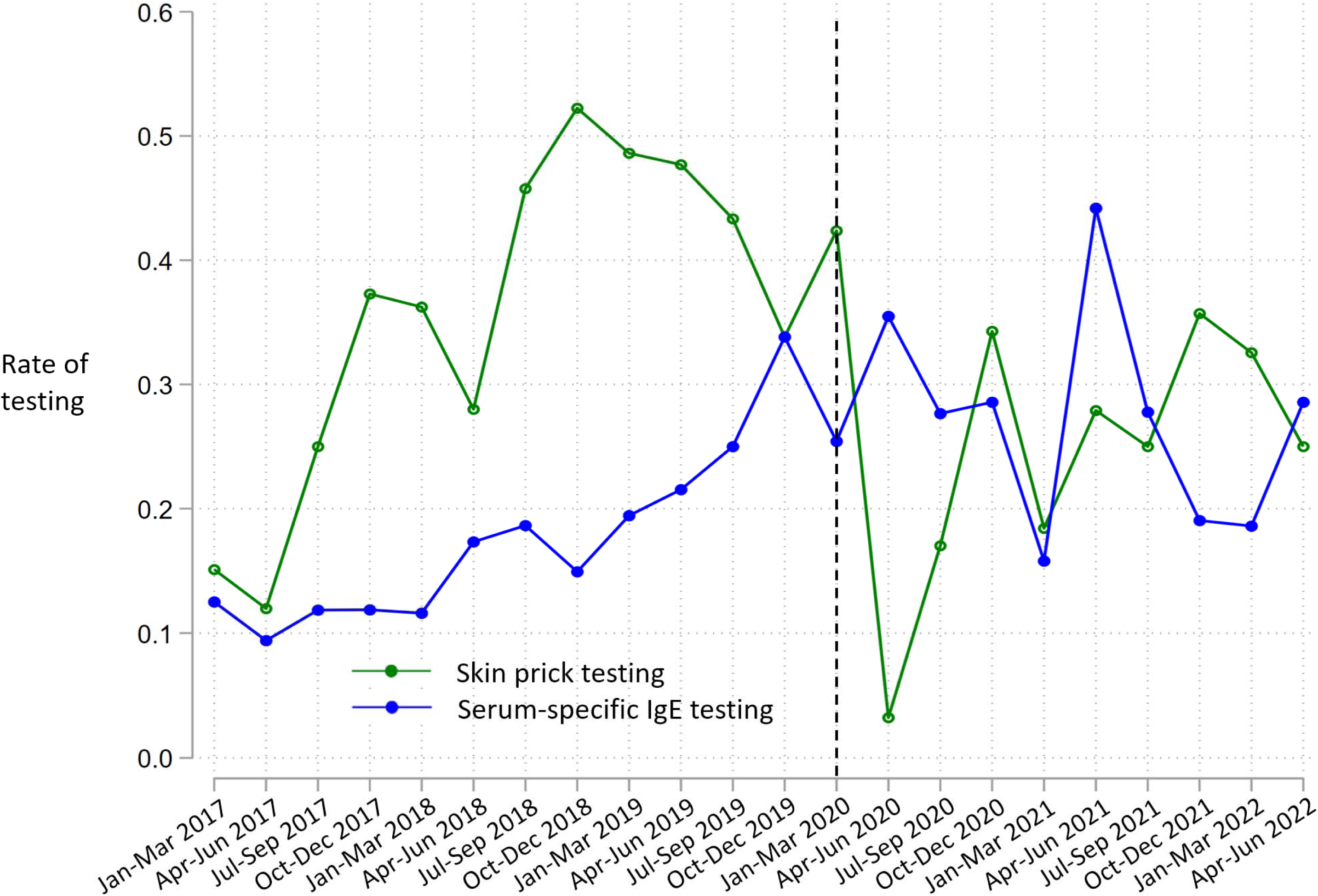
Three-month rates of skin prick and serum-specific IgE tests for 668 patients who had an index visit between 1/1/2017 and 6/30/2022 and received testing at the visit or up to 90 days after. Rates of testing represent ratios of the number of patients tested to the number of index visits within each three-month period. The black dotted line represents the three-month period during which the COVID-19 lockdown in Philadelphia occurred. *IgE*, Immunoglobulin E.

### Receipt of testing was associated with greater benefit among those with asthma who were never smokers and did not have comorbid COPD

Among 1,151/1,383 (83.2%) patients without COPD, 563 (48.9% of 1,141) received testing, and the receipt of testing was associated with fewer OCS bursts (IRR=0.80, p=0.042) [Table 3]. Among 844/1,383 (61.0%) never smokers without COPD, 417 (49.4% of 844) received testing, and the receipt of testing was associated with fewer bursts (IRR=0.68, p=0.004) [Table 3].

### Skin prick but not serum-specific IgE testing was associated with fewer OCS bursts after testing

418/668 (62.6%) tested patients received skin prick testing, of whom 293 (70.1% of 418) received it at the index visit, and 19 (4.5% of 418) also received serum-specific IgE testing within 90 days. 250/668 (37.4%) tested patients received only serum-specific IgE testing, of whom 149 (59.6%) received it within 1 week of the index visit. Compared with the 250/668 (37.4%) patients who received only serum-specific IgE testing, patients who received any skin prick testing were more likely to be under age 45 years (p<0.001), have commercial insurance (p<0.001), be normal weight or overweight (p<0.001), not have COPD (p<0.001), not be prescribed ICS/LABA and LAMA (p=0.04) and have had no OCS bursts in the year prior to the index visit (p=0.006) [Table E2]. According to analyses that employed the three-level testing variable, the receipt of skin prick but not serum-specific IgE testing was associated with fewer bursts (IRR≤0.77 versus no testing, p≤0.02 for skin prick testing versus no testing in each of the primary and secondary analyses) [Table 4]. The postestimation tests showed that skin prick testing was not different than serum-specific IgE testing according to a significance threshold of 0.05.

**Table 4.**
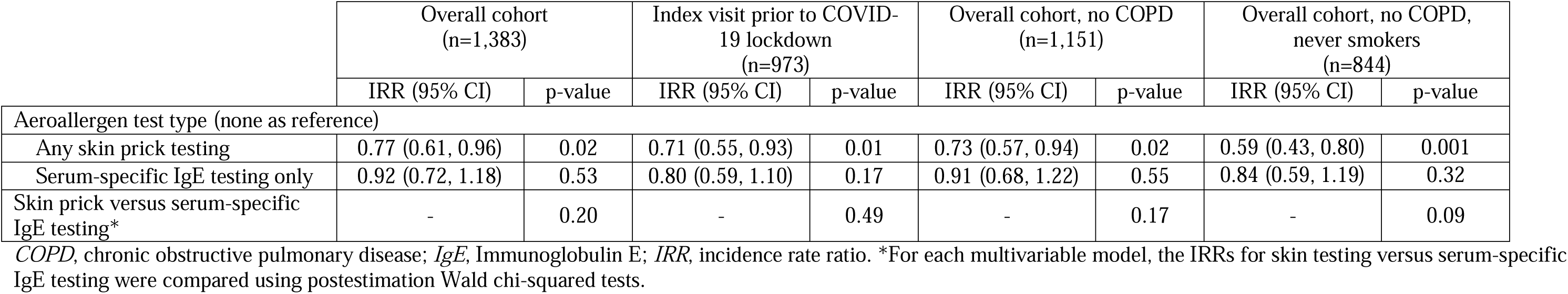
The effect of skin prick testing and serum-specific IgE testing on OCS burst count in the 12-month period after an index Allergy/Immunology visit. The IRRs and p-values were adjusted for all confounding variables in the multivariable models using data from the full cohort of 1,383 adults with asthma who established outpatient care with Allergy/Immunology. This model was repeated for subgroups of the 1,383 patients: 973 whose index visit and post-visit 90-day period occurred prior to the COVID-19 lockdown; 1,151 who did not have COPD; and 844 who did not have COPD and were never smokers.

## DISCUSSION

In this study of real-life adults with asthma in a large health system and who established care in an Allergy/Immunology clinic, and for whom aeroallergen testing appeared to be indicated, the receipt of aeroallergen testing was associated with fewer OCS bursts in the 12-month period after the visit among never smokers without COPD. This finding did not extend to all asthma patients. According to additional analyses, the receipt of skin prick testing had lower IRRs for the outcome of OCS bursts compared with serum-specific IgE testing only, although these effect estimates were not statistically significantly different according to the postestimation tests.

The chart review provided insight into why the clinical guidelines on allergy testing may not be followed, although our findings should be interpreted with caution given the limitations of using EHR data to gather such information. 38% of patients categorized as not tested had prior test results available in the EHR, and these patients’ testing status may be considered misclassified. An alternative view to misclassification is that previously tested patients with uncontrolled asthma or complaints of new asthma triggers could benefit from repeat testing. The U.S. guidelines do not provide guidance in this scenario; thus, some patients who had prior testing may have benefitted from repeat testing. Notably, 20% of patients who underwent chart review had testing ordered or planned by their Allergy/Immunology provider but then testing was not performed.

Our findings suggest that testing in accordance with the guidelines may offer clinical benefit among asthma patients who do not have COPD or a history of smoking. We observed greater reductions in ED visits and hospitalizations for patients who received testing versus those who did not, although we were unable to assess the statistical significance of these trends. It is likely that provider-initiated interventions, patient-driven behavioral changes, and/or greater awareness of asthma triggers are potential mediators between testing and clinical benefit although future studies are needed to understand which of these has the greatest impact on benefit. We previously found that among patients with asthma and positive aeroallergen tests in the same health system, asthma biologic use and allergen immunotherapy were uncommon;^20^ therefore, these therapies were unlikely to account for benefit after testing.

We observed that prior to the COVID-19 lockdown in Philadelphia, skin prick testing was performed at a higher rate than serum-specific IgE testing, but that these rates became more similar after March 2020, presumably due the rise in telemedicine encounters. Prior research has found numerous changes in asthma-related healthcare utilization during the pandemic, including an increase in telemedicine and reductions in asthma exacerbations.^33–42^ We performed a secondary analysis restricting the index visit dates to those that occurred prior to the lockdown and observed greater benefit from testing than in the overall cohort. This difference may reflect a differential benefit according to test type, as patients whose index visits occurred prior to the lockdown received skin prick testing at a higher rate.

Asthma that is associated with comorbid COPD or smoking represent difficult-to-treat subtypes with distinct patterns of airway inflammation and airway remodeling.^29,30^ Our results may reflect the fact that aeroallergen testing and test-based management strategies are less effective among patients with both asthma and COPD. Alternatively, as patients with COPD or smoking histories were older, and older adults with asthma are less likely to be sensitized to aeroallergens,^20,21,31^ older age may underlie their relative lack of benefit from testing. Prospective studies are needed to clarify the relationships among patient demographic factors, comorbidities, aeroallergen test results, and clinical benefit from testing.

Allergy skin prick testing may be more beneficial than serum-specific IgE testing because skin prick testing offers visual results that can be incorporated into management plans at the same visit. One study found that patients prefer test results that can be visualized, such as x-rays and printouts.^32^ By contrast, serum-specific IgE results are usually communicated to patients via telephone or health portal. However, a caveat of comparing the two test modalities in this study is that skin-and serum-tested patients had different characteristics. Specifically, serum-tested patients were more likely to have COPD, be prescribed ICS/LABA and LAMA, and have had more OCS bursts prior to the index visit [Table E2], which were each associated with more bursts after the index visit [Table E3]. The reason for these patient differences in test selection are unknown, although for patients with more severe or uncontrolled asthma, providers may deprioritize a 20-minute skin test procedure in favor of other tests at the visit (e.g., spirometry) or choose to bundle serum-specific IgE testing with other serologic tests. Although we adjusted for confounders in the multivariable analyses, it is possible that residual confounding related to patient selection accounted for the observed differences in the outcome of OCS bursts according to test type. Specifically, we were unable to account for asthma control, environmental exposures, and genetic risk factors for asthma exacerbations. Alternatively, if skin prick testing was more beneficial than serum-specific IgE testing, then our data suggest a disparity in test type selection because patients at higher risk for exacerbations received a less efficacious test type. More research is needed to understand the reasons for this difference in test selection and the relative benefits of skin prick versus serum-specific IgE testing.

This study has several additional limitations. Persistent asthma is a clinical diagnosis, but we used ICS prescriptions to classify asthma as persistent, which may misclassify asthma that has been over-or under-treated. However, as misclassification usually biases toward the null, our results suggest a benefit of aeroallergen testing among patients in the cohort who were correctly classified as having persistent asthma and thus should have received testing according to the guidelines. Our results may not be generalizable to other health systems. However, our large, diverse cohort of adults with asthma in a region disproportionately affected by asthma is a population underrepresented in asthma studies, and therefore, our results may extend to similar health systems. Tested and untested patients may have differed in important, unmeasured ways. For example, it is possible that patients who received aeroallergen testing were more likely to have other types of tests performed, and perhaps the collective benefit of several types of tests resulted in improved outcomes. Due to the study design, patients could have had OCS bursts after the index visit but before testing. However, as most patients who received testing had the testing on or within 1 week of the index visit, such occurrences were minimized.

In summary, we found that in our diverse health system where asthma morbidity is high and asthma disparities exist, patients who received aeroallergen testing in the context of an outpatient Allergy/Immunology visit trended towards fewer OCS bursts in the 12-month period after testing, and this effect was statistically significant for never smokers without COPD and for those who received skin prick testing.

### Abbreviations/Acronyms

BMI: Body mass index
COPD: Chronic obstructive pulmonary disease
ED: Emergency Department
EHR: Electronic health record
ICD: International Classification of Diseases
ICS: Inhaled corticosteroid
IgE: Immunoglobulin E
IRR: Incidence rate ratio
LABA: Long-acting beta-agonist
LAMA: Long-acting muscarinic antagonist
NHLBI: National Heart, Lung, And Blood Institute
OCS: Oral corticosteroid
OR: Odds ratio
VIF: Variance inflation factor

## Data Availability

All data produced in the present study are available upon reasonable request to the authors.

## ONLINE REPOSITORY TEXT

### METHODS

#### Asthma exacerbation measures

An OCS burst was defined as a new prescription for prednisone, methylprednisolone, dexamethasone, or prednisolone occurring at least seven days prior to any other such prescription. Two or more OCS prescriptions within any seven-day period were counted as one burst. Asthma ED visits and hospitalizations were ED or inpatient encounters with any ICD-10 code of asthma, documentation of an oral or intravenous corticosteroid given during the encounter or prescribed upon discharge, plus either 1) a primary ICD-10 code of asthma or a respiratory problem, or 2) a chief complaint of a respiratory problem.

#### Aeroallergen test data

Skin prick test data were available for all skin tests performed at Penn’s primary Allergy/Immunology clinic, where the index visits also occurred. Serum-specific IgE test data were available for tests ordered by any provider and then performed by an in-house or non-Penn laboratory (e.g., LabCorp or Quest) and resulted as structured data (i.e., available under laboratory results in the EHR) in units of International Units/mL.

#### Documentation of the reasons that aeroallergen testing was not performed

We identified seven a priori reasons for why testing was not performed: 1) testing already performed (e.g., at Penn prior to 1/1/2015 or at a non-Penn facility) with results in the chart, and for these patients, the duration from testing to the index visit was categorized into greater than 5 years, 2-5 years, or less than 2 years prior to the index visit; 2) testing ordered or planned by a Penn Allergy/Immunology provider but not performed; 3) patient self-report of previous testing; 4) skin prick testing contraindicated according to a provider (e.g., for patients with uncontrolled asthma at the index visit or at any previous visit in which testing may have been considered); 5) patient declined testing for any reason (e.g., due to fear of the skin test procedure or prohibitive cost); 6) asthma and/or chronic respiratory disease listed as secondary problems in the Allergy/Immunology notes (i.e., testing may not have been high-priority for the provider); or 7) other or unspecified reason for testing not performed. A second clinician reviewed the same 50 charts to assess the interrater reliability of the data collection. For patients whom the two reviewers disagreed on the cause, the reviewers conferred and sought census on the reason.

## ONLINE REPOSITORY TABLES

**Table E1.**
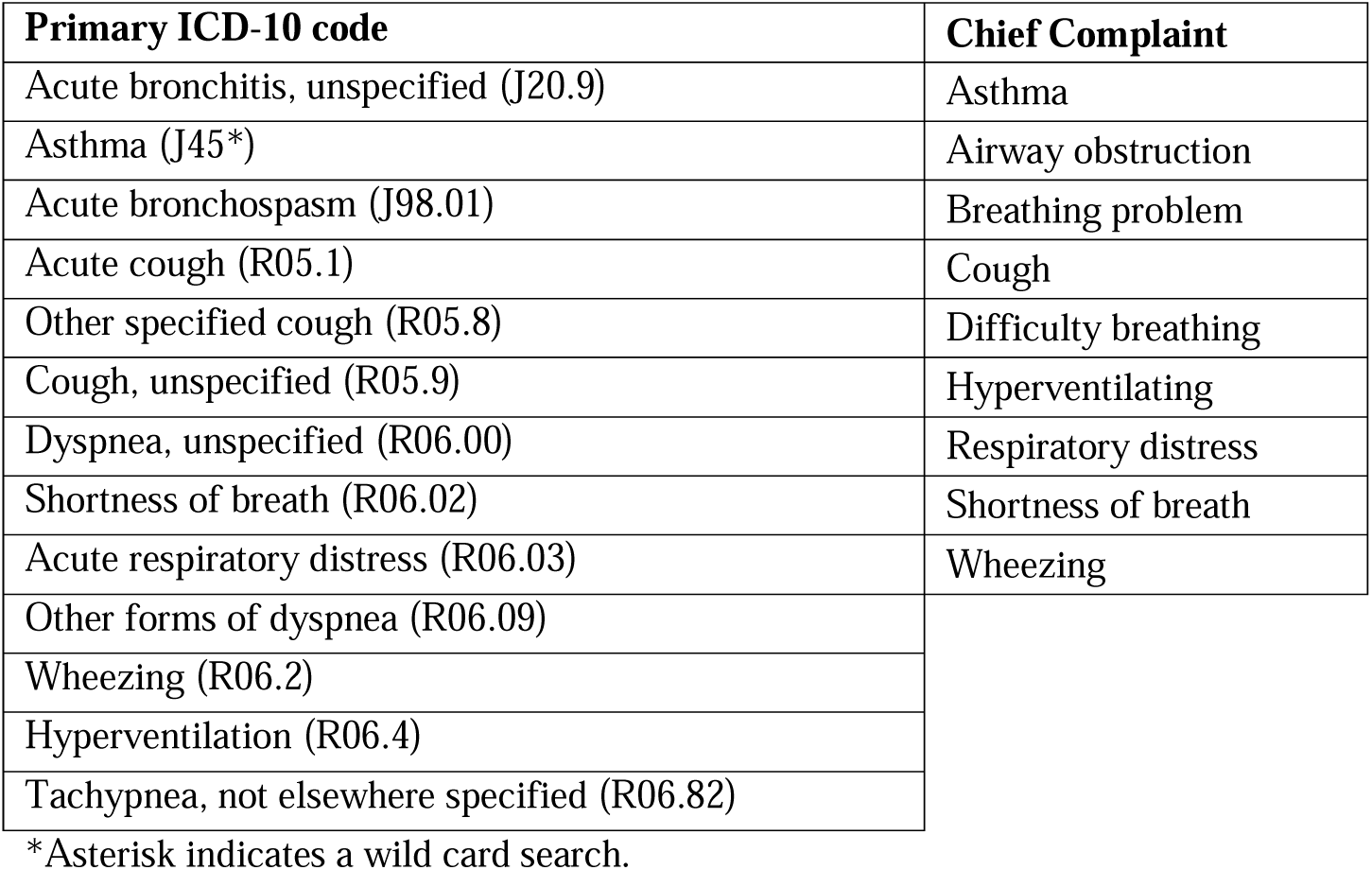
Electronic Health Record criteria used to identify asthma Emergency Department visits and hospitalizations. Encounters were considered to be for asthma if there was any International Classification of Diseases (ICD)-10 diagnosis code for asthma for the encounter, documentation of an oral or intravenous corticosteroid given during the encounter or prescribed upon discharge, plus either a primary code for asthma or a respiratory problem (listed below in alphabetical order by code), or a chief complaint of a respiratory problem (listed below in alphabetical order). The lists of codes and chief complaints were selected from lists of all ICD codes and chief complaints associated with Emergency Department visits and hospitalizations among adults with diagnosis codes for asthma who received outpatient care at Penn.

**Table E2.**
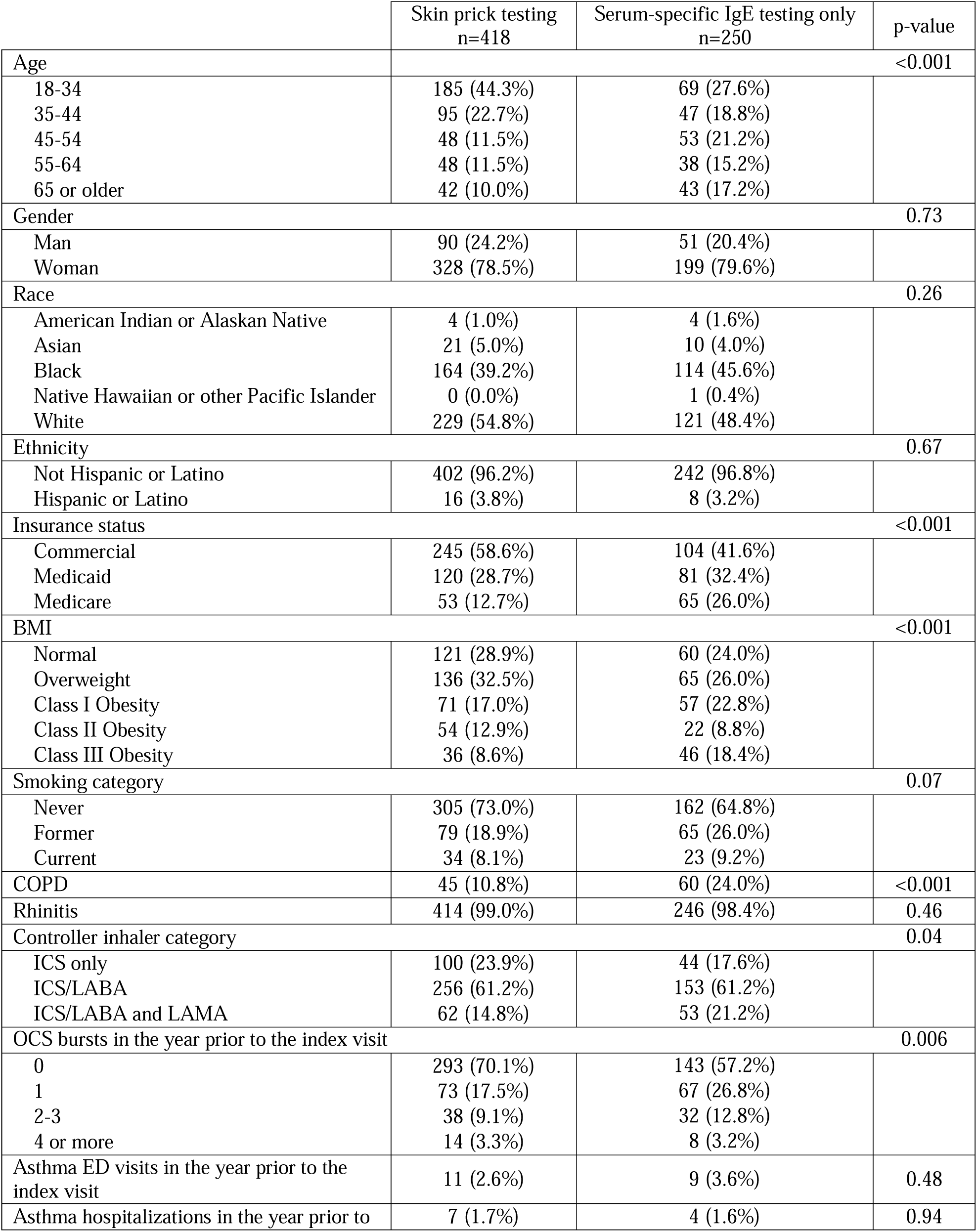
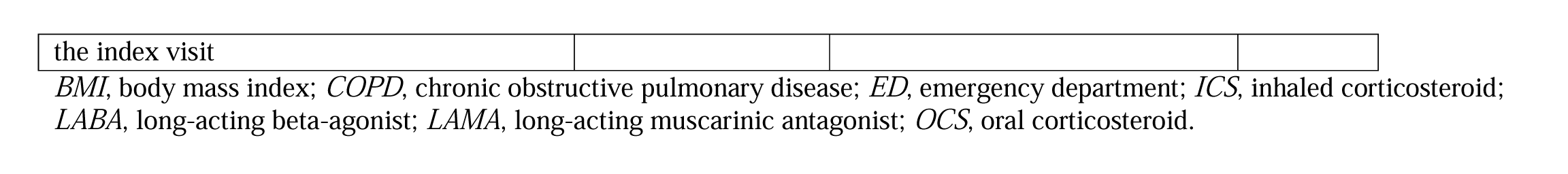
Patient characteristics related to receiving aeroallergen testing among patients who qualified for the study and received testing on or within 90 days of an index Allergy/Immunology visit (n=688). Patients who received any skin testing on or within 90 days of the visit (n=418) were compared with patients who received serum but not skin testing within 90 days of the visit (n=250) using chi-squared tests.

|  | Skin prick testing<br>n=418 | Serum-specific IgE testing only<br>n=250 | p-value |
| --- | --- | --- | --- |
| Age |  |  | <0.001 |
| 18-34 | 185 (44.3%) | 69 (27.6%) |  |
| 35-44 | 95 (22.7%) | 47 (18.8%) |  |
| 45-54 | 48 (11.5%) | 53 (21.2%) |  |
| 55-64 | 48 (11.5%) | 38 (15.2%) |  |
| 65 or older | 42 (10.0%) | 43 (17.2%) |  |
| Gender |  |  | 0.73 |
| Man | 90 (24.2%) | 51 (20.4%) |  |
| Woman | 328 (78.5%) | 199 (79.6%) |  |
| Race |  |  | 0.26 |
| American Indian or Alaskan Native | 4 (1.0%) | 4 (1.6%) |  |
| Asian | 21 (5.0%) | 10 (4.0%) |  |
| Black | 164 (39.2%) | 114 (45.6%) |  |
| Native Hawaiian or other Pacific Islander | 0 (0.0%) | 1 (0.4%) |  |
| White | 229 (54.8%) | 121 (48.4%) |  |
| Ethnicity |  |  | 0.67 |
| Not Hispanic or Latino | 402 (96.2%) | 242 (96.8%) |  |
| Hispanic or Latino | 16 (3.8%) | 8 (3.2%) |  |
| Insurance status |  |  | <0.001 |
| Commercial | 245 (58.6%) | 104 (41.6%) |  |
| Medicaid | 120 (28.7%) | 81 (32.4%) |  |
| Medicare | 53 (12.7%) | 65 (26.0%) |  |
| BMI |  |  | <0.001 |
| Normal | 121 (28.9%) | 60 (24.0%) |  |
| Overweight | 136 (32.5%) | 65 (26.0%) |  |
| Class I Obesity | 71 (17.0%) | 57 (22.8%) |  |
| Class II Obesity | 54 (12.9%) | 22 (8.8%) |  |
| Class III Obesity | 36 (8.6%) | 46 (18.4%) |  |
| Smoking category |  |  | 0.07 |
| Never | 305 (73.0%) | 162 (64.8%) |  |
| Former | 79 (18.9%) | 65 (26.0%) |  |
| Current | 34 (8.1%) | 23 (9.2%) |  |
| COPD | 45 (10.8%) | 60 (24.0%) | <0.001 |
| Rhinitis | 414 (99.0%) | 246 (98.4%) | 0.46 |
| Controller inhaler category |  |  | 0.04 |
| ICS only | 100 (23.9%) | 44 (17.6%) |  |
| ICS/LABA | 256 (61.2%) | 153 (61.2%) |  |
| ICS/LABA and LAMA | 62 (14.8%) | 53 (21.2%) |  |
| OCS bursts in the year prior to the index visit |  |  | 0.006 |
| 0 | 293 (70.1%) | 143 (57.2%) |  |
| 1 | 73 (17.5%) | 67 (26.8%) |  |
| 2-3 | 38 (9.1%) | 32 (12.8%) |  |
| 4 or more | 14 (3.3%) | 8 (3.2%) |  |
| Asthma ED visits in the year prior to the index visit | 11 (2.6%) | 9 (3.6%) | 0.48 |
| Asthma hospitalizations in the year prior to | 7 (1.7%) | 4 (1.6%) | 0.94 |
| the index visit |  |  |  |

*BMI*, body mass index; *COPD*, chronic obstructive pulmonary disease; *ED*, emergency department; *ICS*, inhaled corticosteroid; *LABA*, long-acting beta-agonist; *LAMA*, long-acting muscarinic antagonist; *OCS*, oral corticosteroid.

**Table E3.** Incidence rate ratios (IRRs) and p-values for the remaining independent variables in the primary multivariable model that evaluated the outcome of oral corticosteroid burst count after an index Allergy/Immunology visit for all 1,383 patients.

|  | IRR (95% CI) | p-value |
| --- | --- | --- |
| Age in years (18-34 as reference) |  |  |
| 35-44 | 1.36 (1.03, 1.79) | 0.03 |
| 45-54 | 1.10 (0.82, 1.47) | 0.52 |
| 55-64 | 0.97 (0.70, 1.33) | 0.84 |
| 65 or older | 1.13 (0.76, 1.67) | 0.55 |
| Gender (Man as reference) |  |  |
| Woman | 1.34 (1.05, 1.71) | 0.02 |
| Race (White as reference) |  |  |
| Asian | 0.85 (0.51, 1.41) | 0.52 |
| Black | 0.93 (0.74, 1.16) | 0.52 |
| Other* | 0.98 (0.31, 3.10) | 0.97 |
| Hispanic or Latino Ethnicity | 0.56 (0.27, 1.14) | 0.11 |
| Insurance status (Commercial as reference) |  |  |
| Medicaid | 1.03 (0.80, 1.32) | 0.82 |
| Medicare | 0.92 (0.67, 1.27) | 0.62 |
| BMI (Normal as reference) |  |  |
| Overweight | 1.35 (1.05, 1.75) | 0.02 |
| Class I Obesity | 1.18 (0.88, 1.58) | 0.27 |
| Class II Obesity | 1.43 (1.03, 1.99) | 0.03 |
| Class III Obesity | 1.05 (0.75, 1.47) | 0.80 |
| Smoking category (Never as reference) |  |  |
| Former | 0.94 (0.74, 1.19) | 0.59 |
| Current | 0.89 (0.64, 1.26) | 0.52 |
| COPD | 1.39 (1.08, 1.79) | 0.009 |
| Rhinitis | 1.05 (0.71, 1.55) | 0.82 |
| Controller inhaler category (ICS only as reference) |  |  |
| ICS/LABA | 1.32 (1.01, 1.73) | 0.04 |
| ICS/LABA and LAMA | 2.11 (1.53, 2.92) | <0.0005 |
| OCS bursts in the year prior to the index visit (0 as reference) |  |  |
| 1 | 2.08 (1.66, 2.61) | <0.0005 |
| 2-3 | 3.52 (2.71, 4.56) | <0.0005 |
| 4 or more | 6.29 (4.39, 9.01) | <0.0005 |
| Asthma ED visits in the year prior to the index visit | 1.22 (0.71, 2.09) | 0.47 |
| Asthma hospitalizations in the year prior to the index visit | 1.06 (0.56, 2.00) | 0.85 |

*BMI*, body mass index; *COPD*, chronic obstructive pulmonary disease; *ED*, emergency department; *ICS*, inhaled corticosteroid; *IRR*, incidence rate ratio; *LABA*, long-acting beta-agonist; *LAMA*, long-acting muscarinic antagonist; *OCS*, oral corticosteroid \*The race categories of American Indian or Alaskan Native and Native Hawaiian or other Pacific Islander were combined to form the “other” category due to the small sample sizes.

**Table E4.** Variance inflation factors (VIF) for all independent variables in the primary model of factors associated with oral corticosteroid burst count, based on data from 1,383 patients with asthma.

|  | VIF |
| --- | --- |
| Receipt of aeroallergen testing | 1.06 |
| Age | 1.69 |
| Gender | 1.02 |
| Race | 1.11 |
| Hispanic or Latino Ethnicity | 1.01 |
| Insurance status | 1.60 |
| BMI | 1.08 |
| Smoking category | 1.09 |
| COPD | 1.25 |
| Rhinitis | 1.05 |
| Controller inhaler category | 1.18 |
| OCS bursts in the year prior | 1.09 |
| Asthma ED visits in the year prior to the index visit | 1.08 |
| Asthma hospitalizations in the year prior to the index visit | 1.09 |

*BMI*, body mass index; *COPD*, chronic obstructive pulmonary disease; *ED*, emergency department; *OCS*, oral corticosteroid.

